# *Streptococcus pneumoniae* serotype 38 emerges as one of the dominant serotypes causing invasive pneumococcal disease in Germany and Poland, but not in the Netherlands

**DOI:** 10.1101/2025.03.04.25323220

**Authors:** K. Hajji, I. Wróbel-Pawelczyk, J. van Veldhuizen, K. Maruhn, W. R. Miellet, R. Mariman, A. Steens, N. M. van Sorge, K. Trzciński, M.P.G. van der Linden, A. Skoczyńska, L.J. Visser

**Affiliations:** Centre for Infectious Disease Control, National Institute for Public Health and the Environment - Bilthoven (Netherlands); National Reference Centre for Bacterial Meningitis, Department of Epidemiology and Clinical Microbiology, National Medicines Institute - Warsaw (Poland); German Reference Laboratory for Streptococcus, Department of Medical Microbiology, University Hospital RWTH Aachen - Aachen (Germany); Department of Medical Microbiology and Infection Prevention, Amsterdam UMC, location University of Amsterdam, Netherlands Reference Laboratory for Bacterial Meningitis - Amsterdam (Netherlands); Department of Pediatric Immunology and Infectious Diseases, University Medical Centre Utrecht - Utrecht (Netherlands)

## Abstract

**Introduction:** *Streptococcus pneumoniae* is a major cause of community-acquired pneumonia and invasive diseases. Vaccination prevents pneumococcal disease caused by vaccine serotypes but leads to an increase in disease caused by non-vaccine serotypes.

**Aim:** We aimed to characterise serotype 38 isolates from invasive pneumococcal disease (IPD) patients in Germany, Poland and the Netherlands to explain a recent surge in cases.

**Methods:** IPD surveillance data from 2013/2014 to 2023/2024 were reviewed in all three countries for trends in serotype 38 incidence. Collected isolates were serotyped and phenotypically tested for antimicrobial resistance. Selected serotype 38 isolates (n=130) were sequenced and subjected to phylogenetic analysis, along with 213 publicly available genomes, and characterised according to their virulence and resistance genes.

**Results:** Surveillance data revealed an increase in the percentage of serotype 38 in IPD in Germany and Poland in 2023/2024. This was most pronounced among children aged 0-4 years (from 4.3% to 17.1% and 4.0% to 15.8% of IPD cases, respectively) and adults aged ≥60 years (from 1.5% to 7.0% and from 0.7% to 2.9%, respectively). No such rise in serotype 38 IPD was observed in the Netherlands. Phylogenetic analysis showed that recent isolates mostly emerged from previously circulating strains, showed no significant changes in gene content and carried little antimicrobial resistance genes.

**Conclusion:** The recent surge in non-vaccine serotype 38 among IPD cases in Germany and Poland cannot be explained by changes in antimicrobial resistance or other genetic changes. Our study underscores the importance of international collaboration in monitoring pneumococcal serotype trends, to inform policymakers.

## INTRODUCTION

*Streptococcus pneumoniae* is a major bacterial cause for community-acquired pneumonia and for invasive disease usually manifested by bacteraemic infection and meningitis. The incidence of invasive pneumococcal disease (IPD) is highest among young children and the elderly (1). To reduce the burden of disease caused by *S. pneumoniae*, many countries have introduced pneumococcal conjugate vaccines (PCV) and pneumococcal polysaccharide vaccines (PPV) in their national immunization programs (NIP).

Currently marketed vaccines target 10 to 23 of the 107 known pneumococcal serotypes, which differ based on the chemical composition and immunogenicity of their capsular polysaccharides (2). These vaccines target serotypes with the highest pre-vaccine IPD prevalence. While both PCVs and PPVs prevent vaccine serotype (VT) disease in adults, only PCVs are effective in young children. Since PCVs also prevent VT carriage acquisition, vaccinating children can create herd immunity across a population. However, the introduction of PCVs increased carriage and subsequent IPD cases from non-vaccine serotypes (NVTs), which reduced vaccination benefits (3–5) and prompted higher-valency PCV development.

In Germany, PCVs have been recommended for all children under 2 years of age since 2006. Initially, the seven-valent Prevenar vaccine (PCV7) was used. The 10-valent synflorix (PHiD-CV,PCV10) was licenced in April 2009 and used in parallel with PCV7, and in December 2009 the thirteen-valent Prevenar 13 (PCV13) replaced PCV7. In 2023, the recommendation was changed to vaccination with either PCV13 or the 15-valent Vaxneuvance (PCV15). In Poland, PCV10 was the first pneumococcal vaccine added to the NIP in 2017. Before this, PCV7 and PCV13 were provided free of charge for at-risk children and PCV7, PCV10 and PCV13 through a few municipal programs, and PCVs are still available for private purchase (6,7). In the Netherlands, PCV7 was introduced into the NIP for children in 2006 (8). It was replaced by PCV10 in 2011, and then by PCV15 in September 2024. In addition, all three countries recommend vaccines for older adults. Germany has recently introduced Appexnar (PCV20), while the Netherlands is using Pneumovax 23 (PPV23) at the time of this study. Poland recommends any vaccine suitable for adults, based on the product’s characteristics.

In this study, we describe the rise in IPD cases caused by the NVT serotype 38 in Germany and Poland. To discern whether this increase is due to the spread of a single clone, via recent gene acquisitions or losses, or via changes in anti-microbial resistance (AMR), we conducted genomic analyses on a selection of serotype 38 isolates collected from IPD over the past 10 years in these two countries, as well as in the Netherlands, a neighbouring country where no increase in serotype 38 IPD was observed. Our research highlights the importance of international collaboration in tracking pneumococcal serotype trends and guide policy decisions.

## METHODS

### Data collection

Germany has performed nationwide passive laboratory surveillance for IPD among adults since 1992 and children since 1997. The number of received isolates has increased over time to an estimated 50% of all IPD cases. IPD became a notifiable condition in March 2020, and even though this was not connected to obligatory submission of isolates to the reference laboratory, numbers of received isolates once more increased, to an estimated 80-90%, currently.

In Poland, the passive surveillance system covers the entire country and its population and invasive isolates are collected since 2005. The National Institute of Public Health (NIH) -National Research Institute conducts mandatory IPD surveillance based on notifications from hospitals. Additionally, a voluntary laboratory-based component of surveillance is managed by the National Reference Centre for Bacterial Meningitis (NRCBM) that collects and types pneumococcal isolates from invasive cases.

In the Netherlands, IPD surveillance exists since 2003 through sentinel labs that covered around 25% percent of all labs before the year 2019, and ∼28% after. IPD is notifiable for children born in or after 2006, and since 2021 for adults over 60 years of age (8). IPD isolates are sent to the Netherlands Reference Laboratory for Bacterial Meningitis (NRLBM) at the Amsterdam UMC for typing.

All three countries are using the European IPD case definition (9), and serotyping is done using the co-agglutination and Quellung reactions with antisera for pneumococcal serotypes from the Staten’s Serum Institute. The study covered the 11-year period from 2013/2014 to 2023/2024. The number of IPD cases was reported in epidemiological years (from July 1 to June 30).

### Statistical analysis

The Fisher Exact test was used to test for differences in proportions. Differences were considered statistically significant at *p*<0.05.

### Isolate selection for whole genome sequencing

In total, 136 serotype 38 strains from IPD and the period 2014-2024 were subjected to sequencing (Germany: n=35, Poland: n=71, Netherlands: n=30). In all three countries, isolates were selected to represent different years and various regions.

### Sequences from PathogenWatch

Two-hundred-ninety-one GPSC38 assemblies were downloaded from PathogenWatch on 29 April 2024 (10). After quality control with checkm (11) and panaroo (12), a single assembly was excluded due to contamination, leaving 290 genomes of which 213 were from IPD cases, 65 from carriage and 12 had no diagnosis specified. The selection included 11 assemblies from the Netherlands and three from Poland. The oldest sequence was from 1994 and the most recent from 2018.

### Whole-genome-sequencing and genome assembly

136 isolates subjected to Whole Genome Sequencing (WGS) in this study, 85 (Germany: n=20, Poland: n=35, Netherlands: n=30) were sequenced at the Dutch national institute for public health on a NextSeq550 or NextSeq2000 (2×150bp). An additional 15 isolates from Germany were short-read sequenced on the Illumina MiSeq or Novaseq 6000 platform, and 36 isolates from Poland were sequenced by a commercial vendor (Genomed, Poland) on the Illumina MiSeq platform, resulting in a total number of 136 newly sequenced isolates.

For the 85 isolates sequenced in The Netherlands, DNA was extracted using the automated Maxwell system with the RSC Cultured Cells DNA kit according to the manufacturer’s instructions (Promega, Leiden, Netherlands). During sample preparation, one Dutch sample yielded no DNA after extraction. DNA libraries were prepared using Nextera DNA Flex Library Prep kit (Illumina, San Diego, CA, USA.

Genome assembly on paired-end fastq files was performed using the in-house pipeline juno-assembly v3.0.3 (https://github.com/RIVM-bioinformatics/juno-assembly), which uses SPAdes (13). Assemblies were checked for completeness and contamination with checkm v1.1.3 (11), kraken2 v2.1.3 (14), bracken v2.9 (15), and visualised using MultiQC v1.11 (16). Four assemblies were excluded due to contamination, leaving 131 assemblies.

To obtain a full circular genome of a representative strain, a single isolate was subjected to long-read sequencing using a nanopore MinION with R10.4.1 flowcells, and basecalled and demultiplexed with guppy (v4.2.0, dna_r10.4.1_e8.2_400bps basecalling model). Hybrid assembly for this isolate was done with hybracter v0.7.3 (17).

### Phylogenetics

Recombination corrected maximum likelihood trees were computed separately for the new genomes and the complete dataset using gubbins (18). One further sequenced was excluded from the tree by gubbins due to having less than 70% coverage over the core genome, leaving 130 genomes to be used in all genomic analyses. Genomes of isolates from non-IPD were included while calculating the tree of the complete dataset, but leaves from these isolates were pruned during visualisation with TreeViewer (19).

Multilocus sequence typing (MLST) was performed using the mlst tool from Torsten Seemann and the pubMLST database as accessed on 1 June 2024 (20,21). A core genome of all 130 newly sequenced and 290 previously published GPSC38 *S. pneumoniae* isolates was computed using snippy and snippy-core (22), with the at that time only available full length genome of serotype 38 (GenBank accession: NZ_AP026916).

### Annotation, pangenome and GWAS

Genomes were annotated with bakta v1.9.3 (23). The annotated serotype 38 capsule locus from Bentley et al. 2006 was supplied with the --proteins flag. A pangenome was computed from the bakta output using panaroo v1.5.0 (12), with the strict clean-mode setting. GWAS was performed on the presence and absence of genes from the panaroo output using scoary v1.6.16 (25) as described in Gori et al. 2020. Genes were divided on sampling date into a group before and during/after the year 2020. *P*-values were corrected for false discovery rate using the Benjamini-Hochberg method and considered significant at *p*<0.05 (26).

### Virulence factor and AMR gene analysis

All protein sequences from the virulence factor database VFDB were downloaded on 29 May 2024 (27), and supplemented with *zmpB, pspA* and *pspC* sequences from Croucher et al. 2017 (28). All protein sequences as determined by bakta were blasted with Blast (29) to the prepared VFDB, and the results were filtered with a minimum identity of 90%, a minimum e-value of 1e^-5^, and a minimum bitscore of 200. AMR gene detection was done on the bakta output with AMRFinderPlus and the associated database from 2 May 2024 (30).

### Phenotypic AMR detection

All German isolates were tested for minimal inhibitory concentrations (MIC) of penicillin G, amoxicillin, cefotaxime, erythromycin, clindamycin, tetracycline, levofloxacin, moxifloxacin, trimethoprim/sulfamethoxazole and chloramphenicol using the broth microdilution method as recommended by the CLSI and using the microtiter plates (Sensititre NLMMCS10, TREK Diagnostic Systems Ltd., East Grinstead, UK), and cation adjusted Mueller-Hinton broth (Oxoid, Wesel, Germany) with 5% lysed horse blood.

For Polish isolates, antimicrobial susceptibility to penicillin, cefotaxime, rifampicin, meropenem, chloramphenicol, and vancomycin was tested by the Etest (BioMerieux), the MICEvaluator (Oxoid) or MIC Test Strip (Liofilchem) according to the manufacturers’ instructions. Susceptibility to erythromycin and clindamycin was detected with the disk diffusion method.

The current CLSI and EUCAST criteria were applied for interpretation (31,32).

## RESULTS

### Epidemiology of serotype 38 IPD in Germany, Poland, and the Netherlands between 2013 and 2024

Over an 11-year time span (2013/2014 to 2023/2024), reference laboratories in Germany, Poland, and the Netherlands (sentinel data only) reported n=35,123, n=10,261, and n=6,276 IPD cases, respectively. In Germany and Poland, there has been an increase in surveillance sensitivity, which has resulted in a consistent rise in received IPD isolates over the years, except during the COVID-19 pandemic. Overall, during this period, Germany observed a 2.7-fold and Poland 3.5-fold increase in all reported IPD cases. The Netherlands employed a sentinel lab system covering between 25% to 28% of the country, which consistently reported around 600 cases annually, excluding the COVID-19 pandemic period. Among the pneumococcal isolates sent to the reference centres for IPD, 100% (n=35,123/35,123) in Germany, 97.1% (n=9,960/10,261) in Poland, and 99.4% (n=6,239/6,276) in the Netherlands were serotyped. Of those with identified serotypes, serotype 38 accounted for 2.3% (n=804/35,123) of cases in Germany, 1.3% (132/9,960) in Poland, and 0.7% (n=42/6,239) in the Netherlands. In Germany and Poland, a substantial proportion of serotype 38 IPD cases was reported during the epidemiological year 2023/2024 alone (47.5% or n=382/804 in Germany, 47.0% or n=62/132 in Poland). In both countries it represented a significant rise compared to any previous study year (*p*<0.01 for every comparison). No such increase was observed in the Netherlands. To account for temporal changes in overall incidence of IPD and laboratory efforts, we analysed shifts in proportions of IPD caused by serotype 38 instead of absolute number of cases and investigated possible associations between the rise in this serotype in IPD and age, sex and IPD symptoms.

The surges in serotype 38 IPD cases in Germany and Poland were primarily associated with isolates obtained from children under five years of age and the population aged ≥60 years (Figure 1, Supplementary Table 1). In Germany, the proportion of serotype 38 IPD cases among all children under the age of 5 years increased significantly, from 4.3% (n=61/1418 cases reported during the period July 2013-June 2023, range: 0% to 7.5% yearly) to 17.1% (n=42/245 cases reported in 2023/2024 alone, *p*<0.001). Likewise, in Poland, this proportion increased from 4.0% (n=25/624, range: 0% to 12.2% yearly) to 15.8% (n=18/114) during the same timeframe (*p*<0.001) (Supplementary Table 1). This shift has positioned serotype 38 as the second and the third most common serotype among children under five in Germany and Poland, respectively. It is also important to note that in Poland, an increase in serotype 38 cases to 12.2% was already observed in the 2019/2020 period, prior to the COVID-19 pandemic that started in March 2020. In the Netherlands, in sentinel labs, only 1 case from a child <5 years of age with serotype 38 IPD was found in the study period.

**Figure 1.**
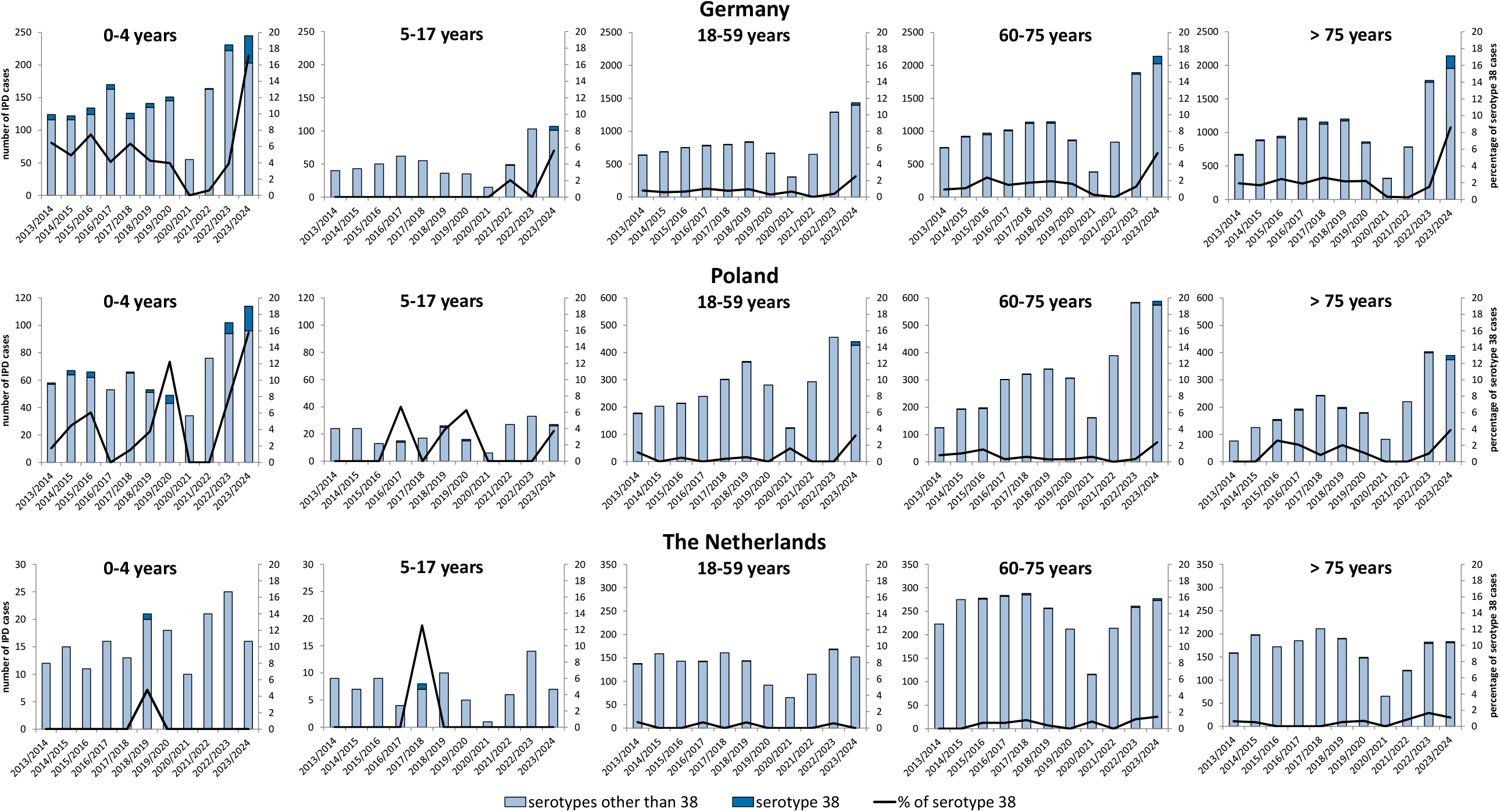
Number of IPD cases with known serotype and percentage of serotype 38 cases in Germany, Poland and the Netherlands over a period of 11 years (from July 2013 to the end of June 2024) divided into age groups.

The rise in the proportion of serotype 38 has also been observed among the elderly (Figure 1, Supplementary Table 1). In Germany, from July 2013 to the end of June 2023, serotype 38 IPD accounted for 1.4% of cases (n=136/9,924, range: 0% to 2.4% yearly) in the age group of 60-75 years old and 1.8% (n=179/9,807, range: 0.3% to 2.6% yearly) in those aged over 75 years. In the 2023/2024, these numbers surged to 5.3% (n=114/2,140, *p*<0.001) and 8.6% (n=184/2,140, *p*<0.001), respectively. Similarly, in Poland, a significant spike was noted from 0.5% (n=14/2,923, range: 0% to 1.5% yearly) and 1.1% (n=20/1,876, range: 0% to 2.6% yearly) in the 60-75 years old and over 75 years of age groups between July 2013 and June 2023, to 2.4% (n=14/588) and 3.9% (n=15/389) in 2023/2024, respectively (*p*<0.001 for both) (Supplementary Table 1). These increases in IPD cases due to serotype 38 has positioned it as the fifth in 60-75 years old and third in >75 years of age in Germany and the ninth most common serotype in Poland for both age groups. In the Netherlands, the proportion of serotype 38 IPD cases among elderly did not change significantly (Supplementary Table 1).

Given the notable rise in serotype 38 IPD cases in Germany and Poland, we conducted an analysis to determine whether the distribution by age, sex or the IPD symptoms presentation have shifted over time. Neither in Germany nor in Poland the rise in serotype 38 IPD was associated with either of sexes or age groups (Supplementary Table 2). In both these countries, sepsis (reported alone or with other presentations) was the most common specified IPD form. In Germany, sepsis accounted for 28.0% (n=118/422) in 2013-2023 and 30.9% (n=118/382, *p*>0.05) in 2023/2024. In Poland, a significant increase of sepsis as a diagnosis was observed from 37.1% (n=26/70) to 58.1% (n=36/62, *p*=0.023) comparing the same periods. However, 42.9% of Polish cases were unspecified in 2013-2023, which fell to 19.4% in 2023/2024, therefore potentially introducing the bias into the results (Supplementary Table 2).

### Phylogenetic analysis with publicly available genomes

To examine potential causes of the recent rise in serotype 38 IPD, 136 isolates from the period 2014-2024 were selected from Germany (n=35, 2018-2024), Poland (n=71, 2014-2024), and the Netherlands (n=30, 2017-2024). Isolates were selected from different patient age groups (0-4 years old: n=64, 5-17: n=3, 18-59: n=9, 60-75: n=25, >75: n=35). After sequencing and quality control, 130 pneumococcal genomes were subjected to genomic and phylogenetic analysis.

All the isolates sequenced in this study were characterised as belonging to Global Pneumococcal Sequencing Cluster 38 (GPSC38) of which 125/130 belong to sequence type 393 (Figure 2). A core SNP phylogenetic tree was built including all 213 GPSC38 genomes from IPD isolates available in PathogenWatch (22). Although some branches appear to have exclusively recent isolates, most of the recent isolates clustered within 25 SNPs with some previously published GPSC38 isolate (Figure 2). Furthermore, there is a clear geographic clustering, with some branches containing samples from a single country with varying sampling dates. No cases were found where the capsular genes of serotype 38 appear in a non-GPSC38 strain (capsule switching), and no changes of note in the capsule operon were found.

**Figure 2.**
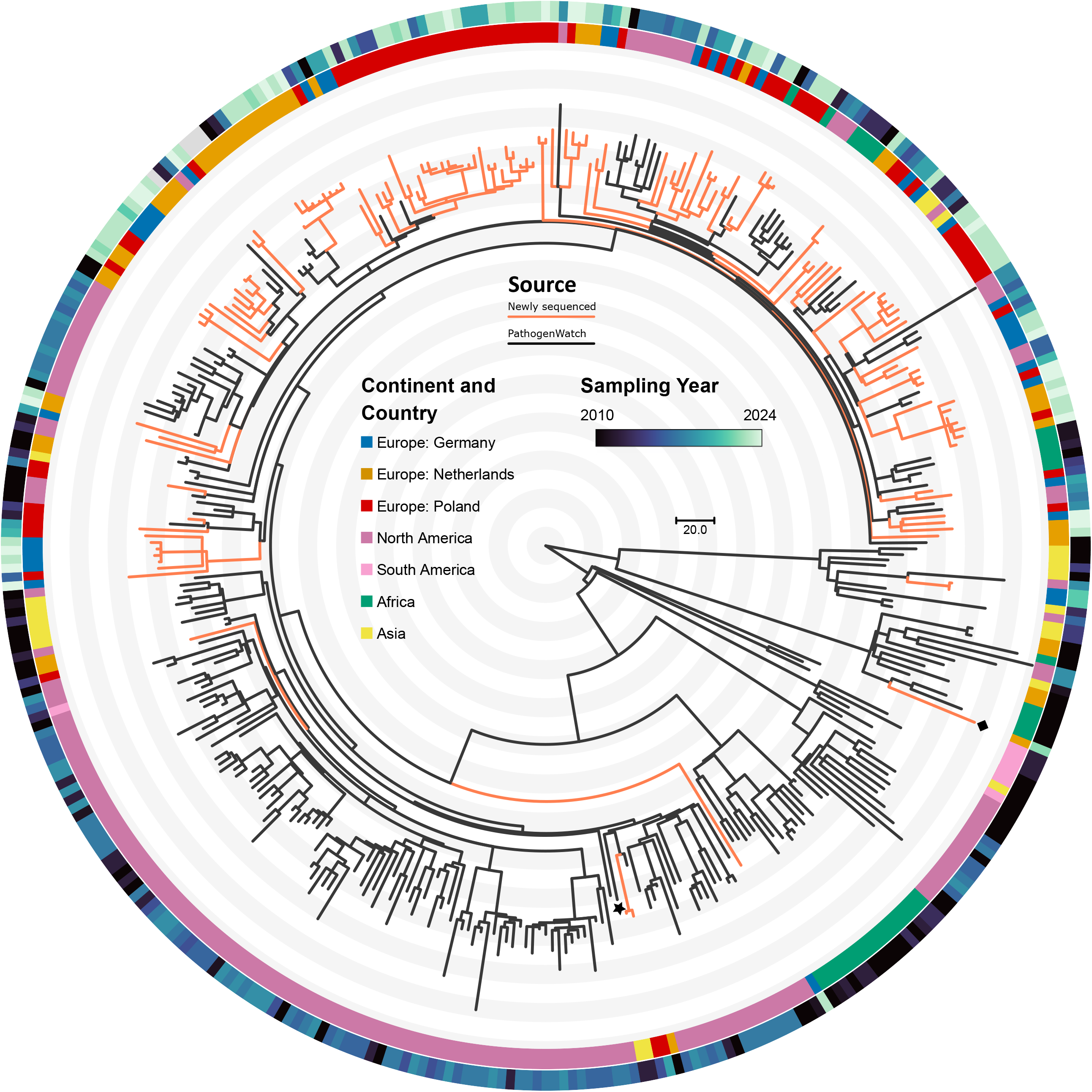
Core SNP phylogeny of 213 publicly available and 130 newly sequenced serotype 38, GPSC38 genomes from IPD. The core genome was determined using all newly sequenced genomes, all GPSC38 genomes found on PathogenWatch, and the only full GPSC38 genome deposited in GenBank (isolate from Japan marked with a black star). A recombination corrected maximum likelihood phylogeny was computed. The new fully sequenced genome is marked with a black square. The scale bar represents the number of core genome SNPs. IPD: Invasive Pneumococcal Disease, SNP: Single Nucleotide Polymorphism. Orange branches represent isolates collected in this study, black branches from sequences from the public domain.

To determine whether recent gene acquisition or loss contributed to the rise in serotype 38 IPD cases in Germany and Poland, a genome wide association study (GWAS) was conducted on isolates sequenced in this study. After computing the pan-genome of the IPD isolates, the presence and absence of genes were compared before (n=40) and after 2020 (n=90). With false discovery rate correction, no significant increase or decrease of genes was found.

### Virulence profile

To characterise the accessory gene content of the isolates in this study, we identified the presence of known virulence factors (Figure 3). All pneumococcal virulence factors present in VFDB were found, except for *zmpC*. All 130 serotype 38 isolates sequenced in this study carried the genes *cbpG, lmb, pavA, pfbB, pce*/*cbpE, pfbA, plr*/*gapA*, and *slrA* coding for pneumococcal adhesin proteins (Figure 3). The choline binding protein gene *pspC (cbpA)* was detected in 91/130 (70%) of isolates. The pilus islet with pilus subunit genes *rrgA, rrgB* and *rrgC* and sortases *srtB, srtC* and *srtD* was found in a single Dutch isolate (0.77%). The IgA1 protease gene *iga* was not detected in this genome, and it was phylogenetically distant from the other isolates. All 130 isolates carried the exoenzyme genes *cppA, eno, degP (htrA), hysA, lytA, nanA, srtA, piuA, psaA, ropA (tig)* and *zmpB*. The number of copies of *lytA* varied among isolates. The gene *ply* for the secreted pneumolysin was detected in all isolates. Autolysin *cbpD* was found in 125/130 (96%) isolates. Pneumococcal surface protein *pspA* was detected in 128/130 (98%) of isolates. Finally, the pneumococcal iron acquisition genes *piaA* and *piaB* were found in 122/130 (94%) of isolates.

**Figure 3.**
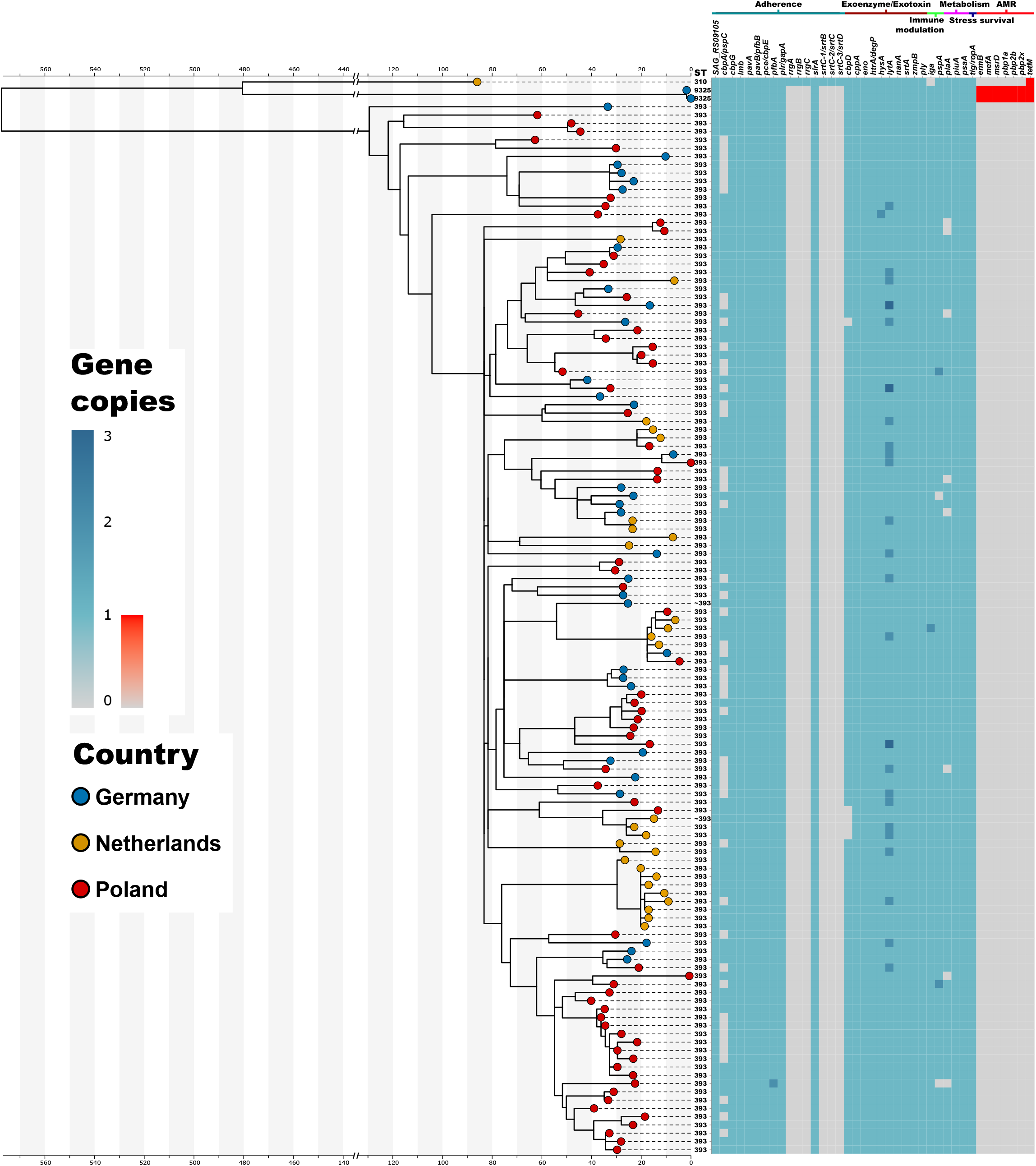
Core SNP phylogeny, virulence profile and antimicrobial profile of the 130 serotype 38 genomes determined in this study. Grey-to-blue scale represents copy number of virulence factor genes while grey-to-red scale represents presence of AMR genes. Genes are sorted on category and alphabetically. Scale bar represents recombination corrected SNP distance. ST: sequence type, AMR: anti-microbial resistance, SNP: single nucleotide polymorphism.

### Antimicrobial resistance

The tetracycline resistance gene *tetM* was found in 3/130 (2.3%) isolates (Figure 3), two from Germany and one from the Netherlands. The two German isolates also carried the *ermB* gene associated with resistance to macrolides and lincosamides, *mefA* and *msrD* associated with resistance to macrolides, and were confirmed to be phenotypically resistant to trimethoprim/sulfamethoxazole. Penicillin resistance was also exclusive to these two isolates and was associated with penicillin binding protein genes *pbp1A, pbp2B* and *pbp2x* allelic type 150:1:77. We did not detect AMR genes in the remaining 128 (98%) isolates and all isolates for which phenotypic data were available were susceptible to all tested antibiotics. No quinolone resistance genes and motifs were found, and all tested isolates were susceptible to levofloxacin and moxifloxacin.

A characterisation of resistance genes in publicly available GPSC38 genomes showed more variation in antimicrobial resistance (Figure 4). The two multidrug resistant isolates described in the previous paragraph cluster with a USA isolate with the same resistance gene profile. These isolates are part of a broader cluster (consisting of MLST sequence types 9325 and 6355) which also contains isolates carrying *ermB*. This broader analysis also reveals four Peruvian isolates that lack *tetM* but do have *pbp2x* allele 184 that is associated with diminished susceptibility to penicillin. However, additional phenotypic data found on pubMLST show that three other isolates from Peru with the same sequence type 5475 and *pbp* type are susceptible to penicillin (21).

**Figure 4.**
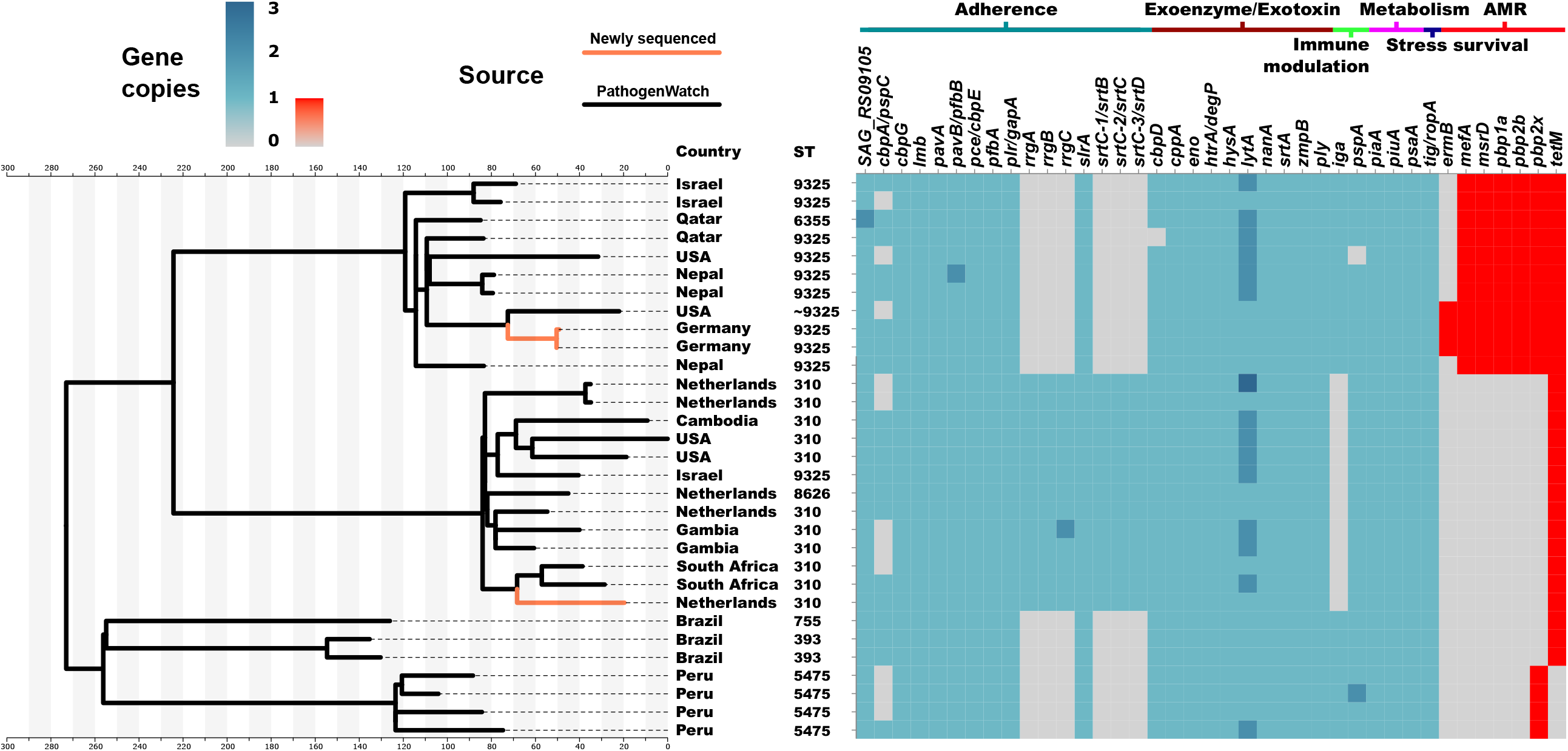
Core SNP phylogeny, virulence profile and antimicrobial profile of resistant branches of n=421 genomes of GPSC38 isolates either sequenced in this study (n=130) or from PathogenWatch (n=291). ST: Sequence Type; AMR: anti-microbial resistance.

## DISCUSSION

In this study, we report on the surveillance of IPD caused by serotype 38 in three European countries: Germany, Poland, and the Netherlands. In recent years, the relative contribution to IPD by this serotype has increased significantly in Germany and Poland, making it one of the top-ranking non-vaccine types. It is important to note that the COVID-19 pandemic most likely stopped the earlier increase in serotype 38 infections in Poland, as indicated by the substantial increase in the number of strains of this serotype in 2019/2020, just before the pandemic. Phylogenetic analysis showed that recent isolates did not belong to a single branch within GPSC38 but were relatively diverse. Expanding the analysis by including published genomes from serotype 38 IPD isolates collected between 1998 and 2018 in 11 countries across all continents, revealed that most of the recent isolates are emergent from variants previously circulating in the respective countries. We therefore exclude that the increase of serotype 38 IPD is due to clonal spread of a single *S. pneumoniae* strain. Analysis of the genomes did not reveal any gene loss or gain, new virulence profiles, or altered AMR characteristics in recent isolates that could explain the rise in IPD caused by this serotype.

Replacement of VTs by NVTs after vaccine introduction has been described extensively. Some research describes these trends in an aggregate manner with NVTs versus VTs (4). In other studies, specific serotypes were associated with vaccine replacement, such as the increase of IPD caused by serotype 19A isolates after PCV7 introduction (3,33,34) and by serotype 24F isolates after roll-out of higher valency PCV13 (35). Despite low incidence in IPD and low prevalence in carriage, serotype 38 was one of the NVTs found to be associated with PCV13 introduction in multiple geographical locations, including Germany, already in the previous decade (4,36). Although Poland uses PCV10 in their national immunization programme, PCV13 is available on the private market, and 26% and 31% of children born in 2017-2018 and 2018-2019, respectively, received this vaccine (4,7,36). In the Netherlands, PCV15 has only recently succeeded PCV10 in the national immunisation programme, after our study period in September 2024. It is tempting to speculate that the absence of serotype 38 emergence in the Netherlands is explained by the lower valency of the used PCV, which would increase the amount competition with serotype 38 pneumococci with PCV13- and PCV15-serotypes that are not in PCV10, such as 19A. Other possible explanations are that secular trends in population immunity and non-pharmaceutical intervention during the COVID19-pandemic have played a role in serotype distributions in recent years. Therefore, it will be important to monitor trends in IPD caused by serotype 38 in the coming years in different countries with different vaccination policies.

Although this study brings attention to the rise of serotype 38 in IPD in Germany and Poland, it remains unclear what the situation is in carriage. At the time of writing, serotype 38 is one of the most common serotype in IPD in Poland, but the frequency in carriage was relatively low in 2016-2020 (37). Without extensive carriage data, it is hard to determine the relative invasiveness of serotypes. Serotype 38 has been reported both as high invasive (38,39), as well as low invasive (40,41), although in the latter case confidence intervals were large due to low prevalence in carriage. Serotype 38 isolates do show relatively little AMR and no capsule switching has been observed, which are properties that are overrepresented in invasive serotypes (42). Recently, the polysaccharide structure was resolved (43).

It is important to note that *S. pneumoniae* serotype 38 has some distinct characteristics that complicate identification with Quellung, *cpsA* PCR and *piaB* PCR. In a survey of pubMLST while writing this manuscript, we found ST393 isolates (the main serotype 38 sequence type) that were incorrectly typed as serotype 25A. The Quellung antigen reagent for serogroup 25 can show cross-reactivity with serotype 38 isolates, which makes further discrimination with factor sera 25b and 25c necessary, as well as testing with serotype 38 specific antigen reagent (44). PCR detection methods might be problematic for serotype 38, as the CDC Streptococcus laboratory describes *cpsA* (*wzy*) PCR testing to be negative in serogroup 25 and serotype 38 (45). Furthermore, our genomic analysis shows that some serotype 38 isolates lack the iron acquisition gene islet containing *piaB* that is among the targets for *S. pneumoniae* detection with molecular methods in polymicrobial upper airways sample testing for pneumococcal carriage (46). Therefore, when relying solely on molecular methods for the detection of pneumococci using a *piaB* qPCR, there is a risk of missing serotype 38.

The surveillance systems in the studied countries have some limitations. First, not all isolates from IPD are sent to the respective reference laboratories, not all strains are viable upon arrival and some materials are not serotypable. In addition, not all isolates that are collected are subsequently sequenced. Because Dutch IPD surveillance did not yet cover all age groups for the entire country during part of the study period, sentinel data had to be used for the Netherlands to allow for following trends. Since Poland and Germany have no sentinel system with dedicated sentinel labs, absolute increases in IPD in recent years can partly be attributed to enhanced surveillance. Therefore, we compared serotype 38 IPD relative to IPD by all serotypes. Another limitation is that antimicrobial susceptibility data for isolates from the Netherlands were not included in this study. AMR results are obtained in decentralised laboratories with non-uniform methods in the Netherlands. However, 29/30 isolates from the Netherlands belong to sequence type 393, which does not show antimicrobial resistance in isolates for which we have phenotypic data. Finally, our method to detect virulence genes is less sensitive when detecting highly variable genes, explaining the lack of *pspC* detection in a substantial fraction of isolates.

Our study highlights the importance of international comparisons to find trends in IPD incidence by different serotypes. New studies or meta-analysis of already conducted carriage and IPD studies might further elucidate the relative disease potential of serotype 38. Nevertheless, the recent increase in IPD caused by this serotype alone makes it a candidate for inclusion in new vaccine formulations, especially if such increase in invasive disease is also seen in other countries.

## Data Availability

All data produced in the present study are available upon reasonable request to the authors

https://pubmlst.org

https://ebi.ac.uk/ena

## Notes

### Competing Interest Statement

The authors have declared no competing interest.

### Funding Statement

Pneumococcal surveillance in Germany was in part supported by Merck/MSD and Pfizer.
This research received partial support from the Polish Ministry of Health (NPOA Project, and the agreement for the activity of the NRCBM), and the Polish Ministry of Education and Science (MIKROBANK 2 Project), as well as grants No. DS-6/2024 from the National Medicines Institute.
The work was supported by the Dutch Ministry of Health, Welfare and Sport.

### Author Declarations

The study was performed with Streptococcus pneumoniae isolates from Germany resulting from routine microbiological diagnostic procedures as requested by the treating physician. Therefore, an ethical approval was not required. Specimens were anonymized and only data on date of birth, sex, vaccination status, type of specimen, diagnosis and hospital/laboratory where the case was diagnosed were registered. The study was conducted as continuous surveillance of the National Reference Centre for Bacterial Meningitis (NRCBM) and in accordance with the World Health Medical Association 1966 Declaration of Helsinki and the EU rules of Good Clinical Practice. The NRCBM was established by the Ministry of Health in 1997 to monitor invasive infections in Poland, including invasive pneumococcal disease. Currently the NRCBM acts under Act of 5 December 2008 on preventing and combating infections and infectious diseases in humans (Dz. U. 2008 Nr 234 poz. 1570) and therefore Institutional Review Board approval is not required. Ethical statement In accordance with Dutch law, approval from a medical ethics committee was not deemed necessary since cases were not subject to any actions or rules of conduct. Data regarding cases were obtained by use of standard surveillance procedures, and pseudonymised data were used in the study. Informed consent was not obtained, as the collection of data complies with the exceptions for not asking informed consent as formulated in the Dutch Implementation Act General Data Protection Regulation.

